# Genomic Characterization of Dengue Virus Outbreak in 2022 from Pakistan

**DOI:** 10.1101/2022.11.07.22281924

**Authors:** Massab Umair, Zaira Rehman, Syed Adnan Haider, Qasim Ali, Zunera Jamal, Muhammad Ammar, Rabia Hakim, Shaheen Bibi, Rida Sagheer, Muhammad Salman, Aamer Ikram

## Abstract

Pakistan, a dengue endemic country has encountered several outbreaks during the past decade. The current study aimed to explore the serotype and genomic diversity of dengue virus responsible for the 2022 outbreak in Pakistan. During August-October 2022, blood samples (n=436) were collected from dengue patients, among which 64.4% (n=281) were positive based on RT-PCR. A subset of DENV-2 and DENV-1 samples were further subjected to whole-genome sequencing. In terms of gender and age, dengue infection was more prevalent in male patients (62.9%) with more adults (77.5%) being infected. Moreover, serotyping revealed DENV-2 to be most predominant serotype (64%; n=180), followed by DENV-1 (35.2%; n=99) and DENV-3 (0.35%; n=1). Phylogenetic analysis of sequenced samples indicates that all the samples (n=8) belong to the DENV-2 Cosmopolitan genotype, falling within a single monophyletic clade that is closely related to sequences from China and Singapore in 2018. Dengue virus dynamics reported in the current study warrants large scale genomic surveillance in order to better respond to future outbreaks.

## Introduction

In the last few years, the dengue virus has become a serious public health concern for developing countries causing a risk of infection to 3.5 billion people each year [1]. According to the World Health Organization (WHO), the dengue virus infects 50 to 100 million people globally every year[2]. Southeast Asia has the highest prevalence of disease with a fatality rate of 70%, followed by Latin America[3] Furthermore, around 16% of cases are reported in Africa and 14% in Europe annually[4, 5]. In Pakistan, the first documented case of dengue fever occurred in 1994 The Ministry of National Health Services, Regulations, and Coordination (NHSRC) reports that since 1994, there have been 1,46,891 laboratory-confirmed cases nationwide, along with 859 associated deaths[6]. Even though the annual epidemic pattern started in November 2005 in Karachi, limited areas of Pakistan encountered dengue before 2006, with the annual dengue epidemics afflicted Pakistan in 2010[7]. Comparably, the dengue cases in Pakistan increased over 14-fold from 3204 cases in 2018 to 47,120 cases in 2019[8, 9]. Recently in 2022, Pakistan is struggling to contain COVID-19 outbreak and simultaneously putting efforts to combat emerging dengue transmission.

In Pakistan, all the DENV serotypes are reported along with the coinfection cases. A preliminary report highlights a high mortality rate in dengue and COVID-19 co-infected patients[10], which could have serious consequences, particularly in dengue-endemic countries. When co-epidemics affect a big portion of the population, countries with few resources, like Pakistan, find it challenging to control them[11]. Population growth, increasing urbanization, weaker vector-control initiatives[12], limited reporting capacity, and a lack of surveillance have all contributed to the spread of disease in Pakistan over the last few decades. There is dire need to understand the dengue circulating serotype and genotypes to combat timely further future outbreak. Therefore, the study aimed to explore the genetic diversity of dengue viruses responsible for the 2022 outbreak in Pakistan.

## Methodology

### Sample Collection and Real-time PCR

The Department of Virology, National Institute of Health, Islamabad received blood samples of 436 NS1-confirmed dengue patients from different districts across the country. Serum was separated and stored at -80°C. Dengue viral RNA was extracted using QIAamp viral RNA extraction kit (Qiagen, Hilden, Germany) as per the manufacturer’s instruction. A four-plex, serotype specific Real-time TaqMan RT-PCR was carried out to detect the DENV serotype while using the CDC protocol [13]. Briefly, a 20-μL RT-PCR mixture with 2x reaction mix, enzyme mix (SuperScript III One-Step RT-qPCR System with Platinum *Taq* DNA Polymerase; ThermoFisher Scientific, Waltham, MA, USA), DENV serotype specific primers and probes and nuclease-free water was prepared. Finally, 5μl of extracted RNA was added to the RT-qPCR master mix and the reaction was set up in a real-time PCR system. Thermal cycling conditions were as follows: RT step 50 °C for 10 minutes, initial denaturation at 95 °C for 5 minutes and 45 cycles at 95 °C for 15 seconds and at 60 °C for 60 seconds. The fluorescent labeled probes were, FAM for DENV-1, VIC for DENV-2, TEXAS RED for DENV-3 and CY5 for DENV-4.

### Next Generation Sequencing

The samples used in this study were tested for dengue using RT-PCR and selected based on Ct values less than or equal to 30. Briefly, using the NEBNext® Ultra II Directional RNA Library Prep Kit (New England Biolabs, MA, USA), the extracted RNA was processed with RNase-free DNase (Roche, Basel, Switzerland) and prepared for unbiased paired-end sequencing (2×150bp) in accordance with the manufacturer’s instructions. By using Qubit dsDNA HS test kit (Invitrogen, USA), the sequencing libraries were measured on the Qubit 4.0 fluorometer and then pooled appropriately in equimolar concentrations. The DNA 1000 Kit and Agilent Bioanalyzer (Agilent Technologies, CA, USA) were utilized to measure the size of the libraries. Finally, at the Department of Virology, National Institute of Health, Islamabad, Pakistan, the pooled libraries were subjected to sequencing on the Illumina MiSeq platform using sequencing reagent, MiSeq Reagent Kit v2 (300-cycles) (Illumina, CA, USA).

### NGS Data Analysis

The resulting raw NGS reads were analyzed using the FastQC programme [14] and assembled into contigs with the SPAdes programme v3.15.5 using the default parameters [15]. The contigs were compared to the National Center for Biotechnology Information (NCBI) Non-redundant (NR) database to determine which reference sequences were the most similar. Utilizing custom scripts, the reads were iteratively mapped three times to the most similar dengue genome with quality trimming and filtering (Trimmomatic ; trimmomatic PE - phred33 ILLUMINACLIP :adapters-PE.fa:2:30:10 LEADING:3 TRAILING:3 SLIDINGWINDOW: 4:15 MINLEN:50) [16], read alignment (Burrows Wheeler Aligner; BWA-mem) [17], and PCR duplicates removal (PICARD; picard MarkDuplicates) [18]. Using the Geneious Prime software v.2022.2 (threshold = 0%, Assign Quality = total, minimum coverage > 10), the consensus genomes were called [19].

### Phylogenetic Analysis

For phylogenetic analysis, the study sequences were subjected to BLAST search in order to identify the closely related sequences. The globally reported sequences were downloaded from NCBI (https://www.ncbi.nlm.nih.gov/). The multiple sequence alignment was performed by MAFFT[20] and a scaled phylogenetic tree was generated by the Maximum Likelihood (ML) using IQ-Tree[21] software with a bootstrap confidence limit based on 1000 replicates.

## Results

During August-October 2022, 436 dengue NS1-positive blood samples were received at the Department of Virology, NIH from 12 different districts of Pakistan. Among these, 281 (64.4%) were found positive for dengue virus when tested by RT-PCR. Dengue infection was more commonly prevalent in male patients (62.9%) compared to females (37%). The age range of dengue patients was between 0 and 72 years. There were more infected adults (77.5%) compared to adolescents (20.9%) and children (1.4%).

Serotyping analysis showed the detection of three serotypes of dengue i.e., DENV-1, DENV-2 and DENV-3. DENV-2 was the most predominant serotype with 180 cases (64%) followed by DENV-1 with 99 (35.2%) whereas only one case of DENV-3 was reported. The highest number of DENV-1 and DENV-2 cases were reported from Rawalpindi with 42 and 61 cases respectively. While the DENV-3 case was reported from Peshawar (Figure. 1).

**Figure 1:**
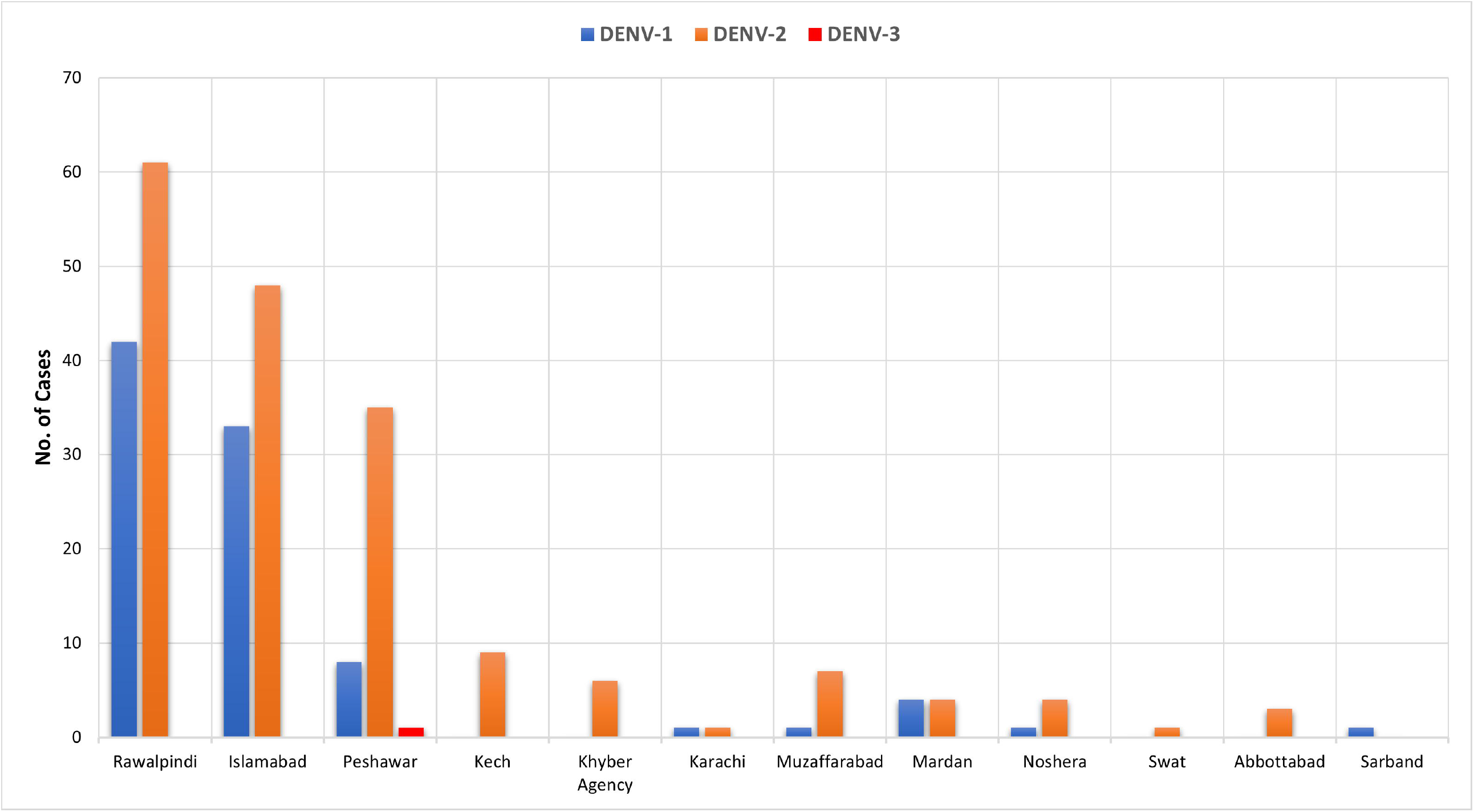
District Wise Distribution of Dengue Serotypes

**Figure 2:**
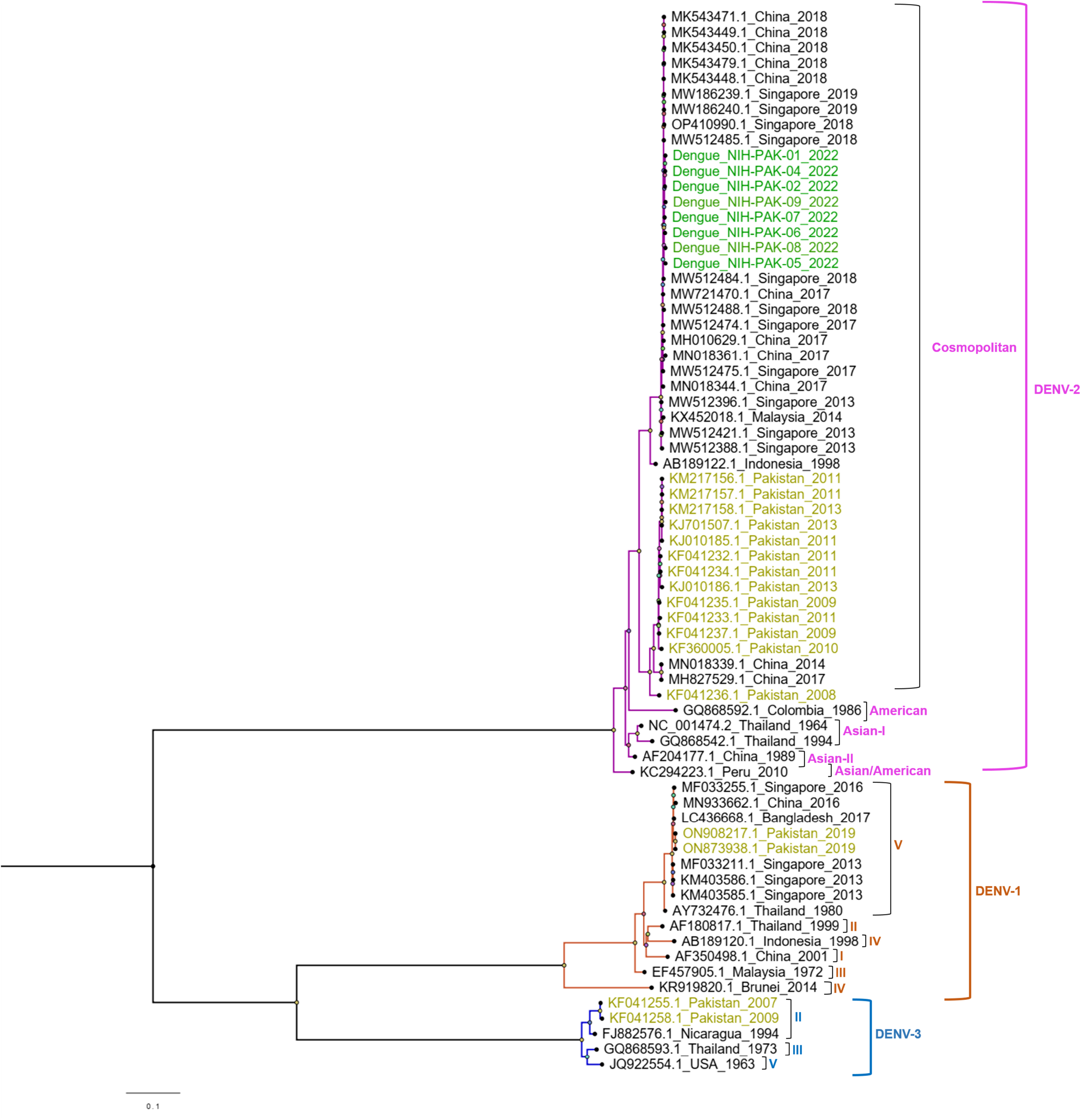
Maximum Likelihood phylogenetic tree of Dengue full genome sequences. The tree was generated through IQ tree with 1000 bootstraps. The study isolates are highlighted in green while the previously reported sequences of DENV from Pakistan are highlighted in yellow.

We carried out whole-genome sequencing of DENV-2 and DENV-1 however, complete genomes of DENV-2 were successfully generated. Phylogenetic analysis of DENV-2 revealed the detection of cosmopolitan genotype. All the study isolates fall within a single monophyletic clade that is closely related to sequences detected from China (MW721470) and Singapore (MW512485, MW512484, and OP410990) in 2018. DENV-2 isolates previously (2008-2013) reported from Pakistan formed a separate cluster. Pakistani DENV-2 isolates from the current outbreak share 99% identity with viruses from China and Singapore, both at the nucleotide and amino acid levels.

## Discussion

In the past few years, Pakistan has encountered several periodic outbreaks of dengue fever with fluctuating severity across various regions. These outbreaks eventually became substantial epidemics in 2011, 2017, 2018, and 2019 [22], with disastrous effects on public health. Wherein, several factors i.e. climate change, inadequate healthcare service, inappropriate vector control, and limited monitoring and surveillance have contributed to the dengue epidemic[23][24].

The current analysis is carried out at a time where Pakistan is experiencing its most catastrophic flood in history where stagnant water has served as breeding environment for mosquito borne diseases including dengue. According to data of National Institute of Health, more than 40,000 cases of dengue have been reported during 2022 majority from flood affected areas. Despite the surge, limited data is available on circulating dengue serotypes and genomic diversity of the viral strains. Therefore, we aimed to explore the serotype and genomic diversity of dengue viruses responsible for the 2022 outbreak in Pakistan.

In line with the overall pattern of circulating dengue serotypes in 2011-2019[22, 25], we found the same pattern in 2022 with DENV-2 being the most prevalent serotype (64%), followed by DENV-1 (35.2%), and DENV-3 (0.35%). However, the trend in Peshawar and Muzaffarabad on circulating serotype reported in 2019 having DENV-1 more than DENV-2 contradicts the current findings. Moreover, an alarmingly high number of cases (46.15%) reported from Rawalpindi contributed to DENV-1. In the case of DENV-3, we reported a single case from Peshawar in 2022, although, dengue infections in the early 1990s in Pakistan were solely due to DENV-3 and continued until 2016. By the end of 2017, a severe dengue outbreak with 100% DENV-2 along with 5% samples having coinfection with DENV-3 was reported in a study[26]. There are several reports on DENV-4 circulating in Pakistan [27, 28], although the present small-scale study does not detect any case in 2022. There have been several reported shifts in dengue virus serotypes, genotypes, or lineages that have resulted in increased severity of dengue infection[11, 25, 26].

Phylogenetic analysis of whole-genomes sequences (n=8) showed the DENV-2 cosmopolitan lineage having a single monophyletic clade, which shares 99% amino-sequence identity with sequences from China and Singapore in 2018. A single monophyletic clade might explain that a similar DENV-2 serotype is circulating in the country. However, the study isolates were distinct from the clade shared by previously reported isolates in 2008 and 2013 from Pakistan. Prior research on DENV-2 dynamics in Pakistan between 2008 and 2013 on partial and complete sequences revealed two distinct clades (IVa, IVb) of Cosmopolitan Genotype IV of DENV-2 strains, in which all the strains from Pakistan were grouped with those from other South Asian nations [29]. Similarly, another study from 2017 reported DENV-2 cosmopolitan genotype (IV), which further bifurcate into sublineages showing close relation with isolates from China, and Singapore [26]. It has been speculated that DENV-2 which remained a dominant serotype in all the major outbreaks in Pakistan might have emerged from two distinct origins i.e. India and Sri Lanka[29]. Nonetheless a more detailed characterization of DENV-2 isolated from Pakistan is required.

## Data Availability

All data produced in the present work are contained in the manuscript.

## Notes

### Competing Interest Statement

The authors have declared no competing interest.

### Funding Statement

This study did not receive any funding.

### Author Declarations

The study was approved by Institutional Review Board of National Institute of Health, Islamabad, Pakistan.

